# Experiences and impact of chronic pain in South Africans living in a rural area: a qualitative study

**DOI:** 10.1101/2025.07.22.25331712

**Authors:** Mamakiri Mulaudzi, Andani Ratshinanga, John Mohale, Tebatso Ramoshai, Michael Evangeli, Tamar Pincus, Romy Parker, Chijioke N. Umunnakwe, Hugo Tempelman, Antonia Wadley

## Abstract

**Objectives:** Previous work in South Africans living with HIV and chronic pain has raised questions regarding maintained levels of physical activity whilst in pain, patterns of pain disclosure and recruitment of social support. Recent data suggest that pain in people living with HIV may be more due to issues of poverty rather than HIV. We explored how South Africans with chronic pain living in a rural area: i) understand and experience chronic pain, ii) how chronic pain affects activity levels and iii) the relationship between pain disclosure and social support.

**Design:** We conducted a qualitative study using in-depth interviews. Transcripts were analysed using thematic data analysis.

**Setting:** The Ndlovu Care Group Research Centre in the rural Elandsdoorn, Dennilton area in Limpopo province, South Africa.

**Participants:** Thirty-four individuals (19 women, 15 men) with mean age 37 years (SD 8) living with chronic pain, half of whom were living with HIV, and half without.

**Results:** Perceived causes of pain included illness or injury, ‘thinking too much’ and non-Western perspectives. Three patterns of activity in response to chronic pain emerged: perseverance, reduced activity, and complete inactivity. Reasons for perseverance included fear of losing income, perceived social stigma, or no social support. Patterns of pain disclosure included full, selective (telling some people but not others depending on their perceived trustworthiness), partial (sharing pain presence but not how severe it was), and non-disclosure. Disclosing pain was common in women and was used to recruit practical support. Men rarely disclosed to recruit support, and if they did, would recruit for financial support. Disclosing pain was also a strategy to avoid the social stigma of being labelled ‘lazy’. Patterns of activity, disclosure, and type of support recruited did not differ between those with and without HIV.

**Conclusions:** Our findings suggest that activity levels, disclosure and recruitment of support in South Africans living with chronic pain are influenced by low income, social stigma, and gender, rather than HIV.

## INTRODUCTION

A national survey of over 10,000 South Africans showed that chronic pain was reported by almost 20% of participants [1]. Our previous work has focused on the experience of chronic pain in South African people living with HIV (PLWH), in whom the prevalence of both acute and chronic pain is more than double that of the general population [2]. Data suggest that the higher pain prevalence in PLWH is likely due to dysregulated immune responses and inflammation, comorbid conditions and infections, and increased prevalence of risk factors such as depression and anxiety, and disrupted sleep [3,4]. More recent data suggest, however, that low socioeconomic status, rather than HIV itself, may associate with the higher prevalence of chronic pain and the risk factors just listed [5,6]. Understanding the experience of pain in low socioeconomic status contexts in South Africa is thus warranted.

One aspect of understanding pain experience is understanding how pain impacts physical activity. Physical activity is typically reduced in people living with chronic pain [7–9] however, using actigraphy, we have found that urban South Africans living with both HIV and chronic pain were as active as their pain-free counterparts [10]. Qualitative comments, in the otherwise quantitative study, suggested that fear of discrimination may have driven individuals to remain active to conceal their HIV status [10]. In a follow up actigraphy study, activity was not associated with HIV stigma [11] and so questions remain about what motivates activity in people living with pain in the South African context.

Another aspect of understanding pain experience is learning how, and to whom, people disclose their pain, and if and how those individuals living with pain ask for support. Preliminary data from the original actigraphy study showed that 44% of participants had not disclosed their pain to friends and 10% had not disclosed to anyone at all [10]. We wondered if this lack of pain disclosure impacted on the recruitment of social support. Understanding patterns of physical activity, pain disclosure and recruitment of social support in people living with chronic pain could help inform pain and symptom management programmes, which are common non-pharmacological solutions to helping people living with chronic pain to manage their pain via exercise and pacing their activity [12].

Pain and symptom management programmes were originally designed in the global north and have been found to be effective there [13,14], but there have been mixed results in rolling out such programmes in South Africa [15–18] and context-specific challenges such as frequent migration for work and HIV stigma may have contributed to those mixed results [17,18]. As such, data on the experience of pain in a South African setting is important to adjust pain and symptom management programmes so they are more context-specific.

We conducted a qualitative study of rural South Africans living with chronic pain. Our aim was to inform pain and symptom management programmes for all South Africans with pain. Our objectives were to learn i) how chronic pain is understood and experienced, ii) how chronic pain affects activity levels and iii) about the relationship between pain disclosure and social support. So we could understand the contributions of HIV to the experience of pain, our cohort was made up half of individuals living with HIV, and half without.

## METHODS

### Study design

This study used a qualitative interpretative research design including in-depth interviews with men and women from rural Limpopo province in South Africa.

### Study Setting

This study was conducted at the Ndlovu Care Group (NCG) Research Centre in the Elandsdoorn, Dennilton area in Limpopo, South Africa. NCG encompasses a research consortium that was conducting a longitudinal study, the Ndlovu Cohort Study, with approximately 2000 men and women, half of whom were living with HIV and half of whom were not. The aim of the longitudinal study was to understand the interaction between HIV and other chronic conditions, with a particular focus on cardiovascular disease [19,20]. This current study was nestled within the larger NCG study.

Elandsdoorn is a rural township in the Moutse area of Limpopo, and is predominantly Black, with the majority of participants speaking IsiZulu and dialects of Sotho, including Sepedi. HIV prevalence in rural Limpopo is 7.4% [21].

### Participants

Individuals attending the NCG Research Centre between April and July 2019 for their clinic visit for the larger research study were invited to take part. Purposive sampling was used: Eligible individuals included HIV-negative and HIV-positive, men and women, aged 18 years and older, with self-reported chronic pain (of all kinds). Chronic pain was defined as pain on most days of the week for a duration of three months or longer [22]. Participants needed to be able to provide written, informed consent and be willing to participate in a face-to-face in-depth interview. Individuals with cognitive impairments were excluded from this study.

### Participant Recruitment Procedure

The nurses and research administrators of the NCG Research Centre assisted with recruitment by identifying individuals reporting pain during their clinic visit for the larger study. Those individuals were invited to participate in the current study and, if interested, were taken to a private consulting room for a full description of the study. Those individuals who met the inclusion criteria and who agreed to participate completed written informed consent and either completed their interview on the same day as their clinic visit or arranged to come back on a separate day.

### Data collection procedure

A semi-structured interview guide (see Supplementary material) was developed to explore questions about participants’ lived experiences with pain (for example, “What do you think is the cause of your pain?”), impact of pain on activity (for example, “When the pain is bad: what are you still able to do?”), disclosure of pain (for example, “Can you tell me about a time you shared living with pain with someone?”) and social support (for example, “Tell me about a time you were able to ask for help when the pain was bad.”). Interviewers were trained on the use of the interview guide and how to probe for further information during interviews. Interviews were conducted by two MSc students (AR and JM), a man and a woman who were able to speak English, isiZulu (AR) and dialects of Sotho including Sepedi (JM). The students had Good Clinical Practice training and some research experience but had not previously conducted qualitative research and so were trained in interview technique by MK, who is an experienced qualitative researcher. Initial interviews were listened to together to refine technique. Interviews were conducted in isiZulu, Sepedi or English based on the participants’ preferred language. Participants who could not speak English or Sepedi had an interpreter to assist. Other than the interpreter on those occasions, no-one else was present during interviews. Interviews were audio recorded for transcription later. Due to impact of experimenter sex on pain reports [23] and to facilitate open sharing of experiences, the male student interviewed male participants and the female student the female participants. In-depth interviews took place in a private consulting room at the Research Facility and lasted between 45 – 60 minutes. No repeat interviews were carried out. The interviewers did not have a relationship with individuals before they were recruited to the study. The interviewers shared that their personal goals for the research were to complete their MSc studies. We planned to recruit approximately 40 people, half living with pain and HIV, and half living with pain without HIV, but with a provision to stop once data saturation was reached.

Prior to in-depth interviews, participants were asked to complete a socio-demographic questionnaire and Brief Pain Inventory. Data were captured through a mobile tablet on REDCap, a secure, web-based software platform designed to support data capture for research studies [24,25] and hosted at the University of the Witwatersrand. The questionnaires were in English and face-to-face interviewer-administered. Interviewers were able to verbally translate questions into the language participants understood if necessary.

Socio-demographic questions included age, home language, highest level of education, employment status and HIV status. Additionally, participants completed the Brief Pain Inventory, a regularly used questionnaire in pain studies, to determine pain severity and interference with everyday activities [26]. Pain severity is determined by asking, on an 11-point Likert scale anchored with 0 ‘no pain’ to 10 ‘the worst pain imaginable’, how bad is their worst, least and average pain, and pain right now. As average pain is not well understood in African contexts where English is not the first language [27], it was excluded and the other three scores divided by three to give an average. The score can range from 0-10. Pain interference asks about pain interference with seven everyday activities on an 11-point Likert scale anchored with 0 ‘no interference’ to 10 ‘complete interference’. The total is divided by seven and so can also range from 0-10.

As part of the larger study, participants had completed the nine-item Patient Health Questionnaire (PHQ-9) to assess the presence of depressive symptoms [28]. Those data were drawn from the larger study database. Nine depressive symptoms are scored from 0 ‘Not at all’ to 3 ‘Nearly every day’ and totalled to give a score, the greatest possible score being 27. A score of 1-4 indicates minimal depression, 5-9 mild depression, 10-14 moderate depression, 15-19 moderately severe depression and >20 severe depression.

### Data Analysis

Thematic data analysis was used to interpret qualitative data following the step-by-step guidelines provided by Braun and Clarke (2006) [29]. The initial analysis of data was conducted by MSc and MA students on the research team (AR, JM and TR) under supervision of MM and AW. MM has extensive training in qualitative research and AW has extensive experience in research on pain and was trained by MM to analyse qualitative data. The initial codes were generated based on the interview guide and the literature review. The data emerging codes were generated from reading through the first four transcripts, engaging in a line-by-line coding. Codes were compared between the students, MM and AW, and disagreements discussed and resolved. Final codes were moved into the next step of analysis by grouping codes to develop themes and subthemes. The themes and subthemes were reviewed and checked against the participants’ direct quotations and approved by MM and AW.

### Ethical considerations

The study was approved by the Human Research Ethics Committee (Medical) from University of the Witwatersrand (clearance number M180652) and permission given to conduct the study in the area by the Limpopo Department of Health. Ethical considerations including voluntary participation, benevolence, confidentiality and anonymity were adhered to. Participants were reimbursed with R200 (£8.50/$11.50) to cover their travel. Participants were allocated a study ID, not known to anyone outside the research team, and this ID is used to refer to participants in the results.

### Patient and public involvement in this study

Neither the study participants nor the public were involved in the design or analysis of this study. Transcripts were not returned to participants for comment and/or correction.

## RESULTS

Data saturation was reached with a total of 34 participants: 19 women and 15 men, 17 men and women were living with both chronic pain and HIV, and 17 were living with pain only. As planned, half of the total cohort (50%, 17/34) were living with HIV. Approximately half of those people living with HIV (53%, 9/17) were virally suppressed (<50 cp/mL). Of all the people asked to take part, 88 women and 99 men declined. Participant characteristics can be found in Table 1.

**Table 1.** Participant socio-demographics.

| **Sociodemographic variables** | **Number (%)** |
| --- | --- |
| **Gender** |  |
| Female | 19 (56) |
| Male | 15 (44) |
| **Age\*** | 37 (8) |
| **Home language** |  |
| isiZulu | 27 (79) |
| Southern Sotho | 4 (12) |
| Northern Sotho | 2 (6) |
| Tsonga | 1 (3) |
| **Highest level of education completed** |  |
| Primary | 1 (3) |
| Secondary | 26 (76) |
| Tertiary | 6 (18) |
| Missing | 1 (3) |
| **Employment** |  |
| Full-time employed | 9 (26) |
| Part-time employed | 7 (21) |
| Unemployed | 18 (53) |
| **Brief Pain Inventory<sup>II</sup>** |  |
| Pain severity score | 4 (1-6) |
| Pain interference score | 5 (0-9) |
| **PHQ-9** |  |
| No depression | 18 (53) |
| Minimal | 7 (21) |
| Mild | 5 (15) |
| Moderate | 0 (0) |
| Moderately severe | 3 (9) |
| Severe | 1 (3) |

\*Mean (SD); <sup>II</sup>Median (Range)

### Qualitative findings

Data analysis yielded seven key themes and twenty-one sub-themes with six sub-subthemes. The following themes will be presented with their subthemes: Participants’ experiences of, and beliefs about chronic pain; participants’ perceptions about causes of chronic pain; the impact of chronic pain on activities; the treatment of pain; reasons for disclosure of chronic pain; patterns of chronic pain disclosure and peoples’ reactions following pain disclosure by participants. See detailed list of themes and subthemes in Table 2.

**Table 2:**
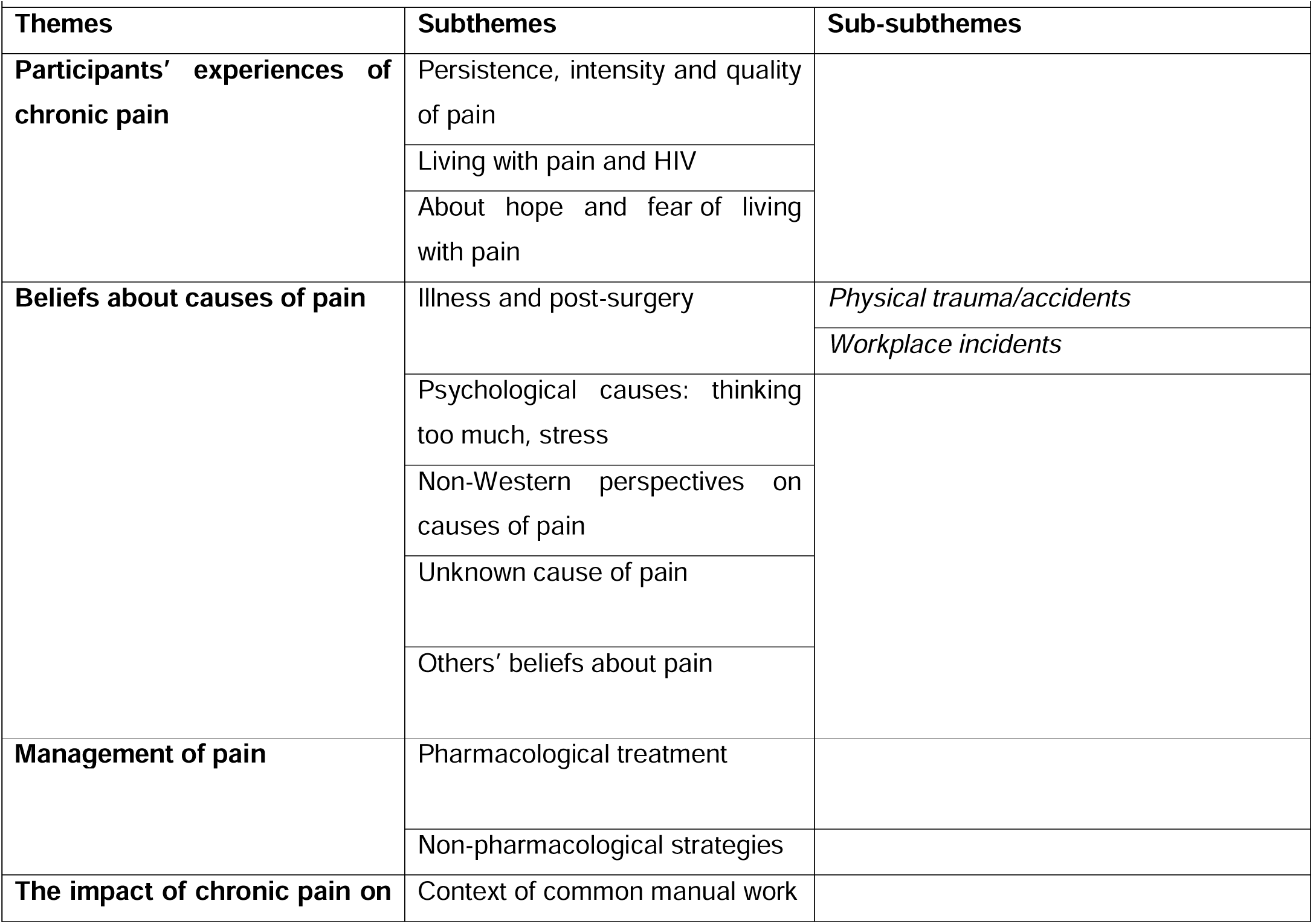
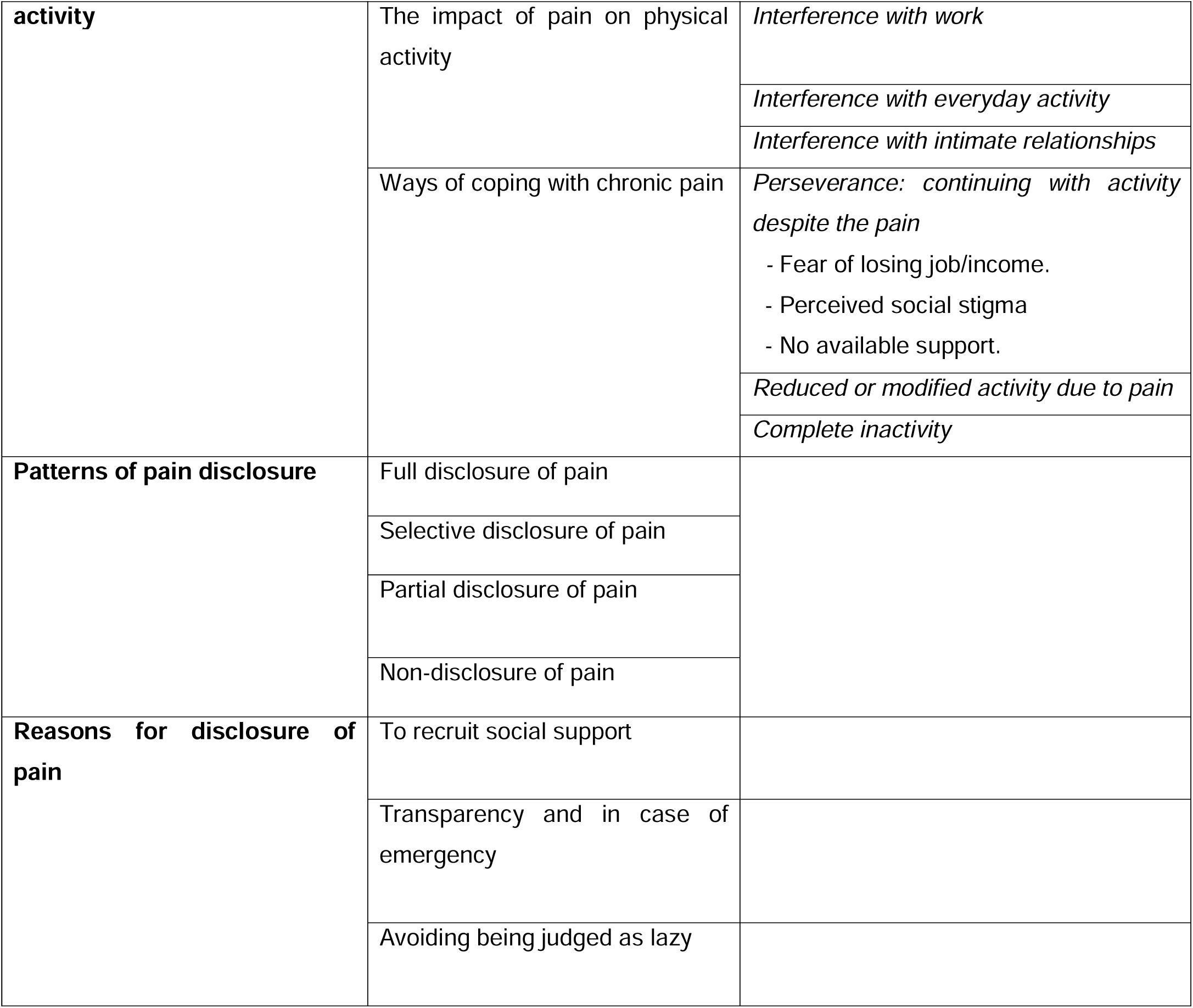
presentation of themes and subthemes.

### Theme 1: Participants’ experiences of, and beliefs about chronic pain

#### Persistence, intensity and quality of pain

Most participants experienced their pain as persistent. Participants described their pain as one that would *‘never completely stop’*. Although the pain could ease and was not felt at every hour of the day, it was something that they perceived as a constant part of their life experience. The intensity of pain could be unpredictable. For example, one woman experienced pain that would ease or subside and then return *‘out of nowhere’* (Participant A083-Female in her 40s). Additionally, she reported that she could feel when the pain was coming. Pain was described frequently by participants both living with and without HIV as “*burning*” or “*stabbing*”.

#### Living with pain and HIV

Many participants living with both HIV and pain commented on their experience of living with both. Most of the participants, particularly men, shared that it was better to live with HIV than pain. For example, two participants shared that:

> “I’ve accepted my status [HIV status] the way it is. So, it’s not something that bothers me anymore, but the pain it’s unbearable because it…reminds me…it just comes and says, “Don’t forget that I’m still here” (**J018, Male-living-with-HIV in his 30s)**

> “HIV I just live with it, but it is not a problem at all, just the pain I have on my back but nothing in connection to HIV, just the pain in my back nothing else.” (**J097, Male-living-with-HIV in his 50s)**

Interestingly, another participant from the interviews with men, expressed a sense of helplessness and sorrow about having to live with both pain and HIV.

> “It’s bad…You feel pain every day, it’s not good… So the freedom of the soul isn’t there. I am not happy about it because I was not born with these things [pain and HIV], I found them here on earth….it doesn’t sit well with me.” (**J030, Male-living-with-HIV in his 50s**)

#### About hope and fear of living with pain

While some of the participants expressed a sense of hope that their pain would be healed, other participants reported a sense of fear about the pain progressing to a more severe state or even fatality. Interestingly, the sense of hope felt amongst some of the participants was recorded in the interviews with women, while a sense of fear was recorded in the interviews with men. Both women and men reporting these emotions were younger than the average of participants in the study.

> “…Because I don’t want it [the pain] to oppress me, because I still have the belief that I will get healed and one day things will go back to normal.” **(A106- Female in her 20s)**

> “That pain is on the lung side. It can affect me in future and I’m still young. If it continues, this kind of pain in the chest… eish, anything can happen.” **(J007- Male in his 30s)**

> “Ja, ja. I’m, I’m afraid, I’m scared. I’m scared that eh, maybe I may damage it more. It can get worse…And then once I damage this backbone, it’s over about my life. I won’t… [I am] no longer going to use my feet to walk.” **(J088- Male in his 20s)**

### Theme 2: Participants’ perceptions about causes of chronic pain

The perceived causes of pain included illness and post-surgery, physical trauma from accidents, and psychological causes. There were also some non-Western perspectives about the causes of pain and for some, the reasons were unknown. None of the participants living with HIV mentioned HIV as a cause or contributor to their pain.

#### Illness and post-surgery

Most participants, especially women, reported that they believed their pain was because of illness or disease, such as TB or a stroke, while others reported that their pain started after surgery including a hysterectomy or after receiving stitches (following a stab wound).

##### Physical trauma/accidents

Several participants, both men and women, believed that their pain was due to an accident or physical trauma. The type of accident was not always probed but when offered was often a road traffic accident.

##### Workplace incident

At least two participants, a woman and a man, reported that they believed their pain was caused by an incident at the workplace. They described their experience as an injury that occurred during work or because of the type of work that they do. For example, the woman participant reported that she had to stand for long hours while working at the time she was employed, whilst the man reported being injured having fallen from scaffolding.

#### Psychological causes: thinking too much, stress

Several participants, both men and women reported a psychological cause of their pain because they were “*thinking too much*” or had “*stress*”. One of the participants expressed food insecurity as the cause of her worry:

> “I think it’s because I don’t have parent[s], I live alone, I don’t work… Ja because I think about these things every morning when I wake up, thinking what am I going to eat? How am I going to make ends meet this month?” **(A106- Female in her 20s)**

Another participant mentioned the combination of working hard and thinking too much.

> “I think that it’s [the pain] caused mostly by thinking too much. Thinking and working, working hard. Here by my heart, sometimes by my head, mostly headaches and mostly if I work too much. I feel it in my body, I get headaches, here my feet, you see.” (**J108-Male in his 30s**)

#### Non-Western perspectives on causes of pain

Some of the women reported perceived causes of pain that do not fit with a Western medical understanding. For example, a middle-aged woman explained:

> “I breathe with my heart. I no longer breathe using my lungs, but I breathe using my heart so that’s why after some time it tires. The pain is caused by the lungs that are malfunctioned and the heart is getting weak.” **(A084 - Woman-living-with-HIV in her 40s)**

An interesting narrative about understanding of pain came from an interview with a woman who shared what sounded like a supernatural understanding of her pain. The participant shared that the pain was due to a genetic predisposition since her parents both had arthritis but this predisposition may have been due to one of her parents being bewitched. The participant further explained that that parent consulted a traditional healer with the aim of protecting their children from inheriting the painful condition.

> “I have arthritis …Well…. actually, ..my parents have arthritis. They told me it’s inherited but I’m not sure. It’s like they were bewitched and then that traditional healer helped my parents so that that thing [pain] won’t come to us.” **(A047 Female in her 30s)**

#### Unknown cause of pain

Most of the women had an idea of what had caused their pain, whereas about half of the men said they did not know. Those men responded with a brief “I don’t know” to the question. One of the male participants elaborated further, however, wondering if his pain could be caused by cold weather. He said: “Eish I don’t know what is causing it now, [if] it’s the cold or what, I don’t know**.*”* (J064 -Male-living-with-HIV in his 40s)**

#### Others’ beliefs about pain

Some of the participants relayed responses from their family about their beliefs about pain. Two narratives from the interviews with women suggested surprise and shock from relatives because of the belief that pain is only for old people.

> “She was surprised, maybe shocked at this age, this pain is for older people. Yah she was shocked and sad” **(A047- Female in her 30s)**

> “My [sibling] was shocked about it because they normally say that the pain at the back affects old people but I am still young.” (**A096-Woman-living-with-HIV in her 40s)**

Another participant touched on an African perspective that influenced beliefs about the cause of pain as originating from malicious intention to harm others through witchcraft. The participant shared that after telling her parent about her pain, the parent was scared that the pain was due to witchcraft. She said that:

> “Yes, she was scared when I explained to her about that. She was scared because with us black people we believe in witchcraft. I think maybe she was afraid of that. I told her there is no one who is witch crafting [bewitching] me” **(A033- Female in her 30s)**

### Theme 3: The impact of chronic pain on activities

This theme focused on the effect of chronic pain on the participants’ activity. The theme is supported by five subthemes including: context of frequent manual work; the impact of pain on physical activity; the impact of pain on intimate relationships; modified activity/reduced household chores due to pain and inactivity due to pain. The impact of chronic pain did not differ between those living with and without HIV.

#### Context of common manual work

Most of the participants engaged in manual work ranging from daily household chores to paid work that included construction or security work or as a cashier. Office administrative work was less common. Some of the manual work described by the participants reflected heavy duty activities such as bricklaying, pushing heavy wheelbarrows all day, handwashing laundry, carrying water or standing for long hours at a job.

> “I do many jobs: I mix, I dig…I have to build for the whole day.” **(J030- Man-living-with-HIV in his 50s)**

> “I prepare water^1^ for the child[ren] because they are going to school….I iron the clothes. I go to the tap (community tap) [to] fetch the water….I clean the house and I cook for them so that after school they can get the food ready.” **(A008- Female in her 30s)**

#### The impact of pain on physical activity

Participants described the negative impact of pain on their daily physical activity. Participants reported how their pain interfered with their work, general everyday activity and intimate relationships.

##### Interference with work

Several of the participants reported how the pain interfered so much with their work that they couldn’t do it anymore. One of the participants who worked as a security guard explained how he had to stop working given the nature of his job and the pain he experienced, and another man reported how he couldn’t brick lay anymore.

> “I was working in security department…When you are working you wouldn’t be allowed to sit because you are having pain. So, I resigned. They didn’t fire me.” (**J097-Man-living-with-HIV in his 50s**)

> “Bricklaying, that’s the hardest job (laughs)… but since uhm I’ve started having pains, I can’t do it anymore because once I do it, eish it’s [the pain is] going to be hectic.” **(J018 -Man-living-with-HIV in his 30s)**

##### Interference with everyday activity

Both men and women described that they “can’t do anything” when the pain is severe. The pain would impact all activity including household chores, and carrying things including children.

> “Eh there’s nothing I am able to do. When I hold something it falls. I’m just sitting. I just sit on the couch busy with my phone [when the] pain is too severe.” **(A084- Woman-living-with-HIV in her 40s)**

> “Yoh when the attack is serious, I cannot do anything. Yoh! I am not able to even stand. I am not able to even sit. I just can’t. [Even] Sleeping, I have to be cautious, ja, I am not even able to hold a baby, to carry anything. I just run out of ideas.” **(A007- Woman-living-with-HIV in her 40s)**

##### Interference with intimate relationships

For two of the women who lived with pain, they shared that the pain had a negative impact on the sexual relationships. One of the women explained that she cannot be in a sexual relationship due to the fear that she might not be able to have sex. The other participant who was in an intimate relationship explained that she couldn’t have sex with her partner due to the pain.

> “No, I avoid, even relationships. It hurts because I would like to have a partner I can share things with and even be happy with… I’m unable to have sex because there are other positions I can’t perform. I am single because of it…I am single because I think I can’t perform those positions in bed, and I feel bad and lonely and small.”**(A047- Female in her 30s)**

> “Like uh, it [the pain] changes, it changes all my mood … I can’t do anything. I can’t even do intercourse with my partner…so, it’s completely interferes.” **(A032- Female in her 40s)**

#### Ways of coping with chronic pain

There were three ways participants had of coping with the pain and painful episodes: Some persevered, some modified or reduced their activities, and others stopped doing anything.

##### Perseverance: continuing with activity despite the pain

Several participants reported instances where they had to continue with work despite feeling the pain for reasons such as fear of losing their job or income, perceived social stigma or no availability of support.

###### Fear of losing job/income

Other participants commented that their perseverance despite pain was fuelled by the fear of losing their job or not getting paid in full for hours not worked due to pain.

> “There is a day at work I couldn’t even pick up the baby. The baby was lying there crying, I was just giving the baby his bottle. I couldn’t do anything to help the baby… And I was afraid to tell his parent about my pain. I thought maybe I would lose my job. Sometimes I think maybe she will go and get someone else, so I continue doing my job while in pain to keep my job. Because that money she’s giving me help me with the food for the kids.” **(A013- Woman-living-with-HIV in her 40s)**

> “No, they will not pay you. Then they tell you that if you [do] not [work]…they can get another one [person]…who’s serious, you are not serious about your work. Where I [am] working [for] a low income.… every second counts…If you [are] off from those seconds, they cut your salary….So, I have to push… to get like reasonable cents….for a better living.” **(A032- Female in her 40s)**

Another participant related his experience to persist working despite the pain to a lack of money for treatment. He explained how he has come to accept that he has to live with the pain:

> “As I’ve said… I’ve experienced this pain … maybe [it] needs serious help … and I’ve experienced it only in the morning. And as I do seek that help, but for now, that help require money… like serious [money]. So, I [have] decided I have to handle this pain, live with it and this pain could stop my life or stop whatever I’m doing. Yes, it’s concerning … it’s part of my health [life], but I can maintain it, for now…. So… I can’t stop working because of it.” **(J007- Male in his 30s)**

###### Perceived social stigma

One of the women participants who is living with pain and HIV, shared that she had thoughts about how people will perceive her if she had to modify her physical activities on the job:

> “I just start wondering how people will start treating me and viewing me, so I just continue with my work and taking my treatment.” **(A096- Woman-living-with-HIV in her 40s)**

###### No available support

Two of the women participants explained that they had to continue engaging in household activities because there was no one available to help them and they had to meet their roles and responsibilities of the household.

> “I’m on my own. My parents are in their 60s and 70s [and] the kids are still young. I cannot say they are going to come back from school and sweep and what about the cooking? If I am in too much pain that’s when my [parent] will come and stand up and ‘let me help you here and there’ but she is old. I’ve just told you that, I mean like, I’m home alone so the boys are like young. I’m the only lady in the house. My [parent] is too old.” **(A089- Woman-living-with-HIV in her 20s)**

> “These chores must be done; no one is going to do them for me…I fetch water.. from outside, and take the water inside. Yes, who’s going to fetch water for you? My kids are young, a child won’t be able to carry a 20 litre bucket.” **(A060 -Woman-living-with-HIV in her 30s)**

#### Reduced or modified activity due to pain

Several participants modified their activities because of the pain, and as a way to manage it. Most of those participants shared that they had cut down on, or modified, the household chores that they used to do. Those who had family support, were able to reduce their household workload. More women than men reported modifying activities due to pain.

> “…But now that I have this pain I no longer clean the way I used to clean. I even do small amounts of laundry because you can’t be bending the entire time.” **(A060- Woman-living-with-HIV in her 30s)**

> *“*I take a chair and sit down and work like this.” (**J074-Male in his 30s)**

#### Complete inactivity

While most of the participants in this study reported persisting with activity, other participants shared a different experience of living with pain. They shared that in instances where the pain is severe, it can render them physically inactive such that they are unable to do anything but wait for the pain to stop.

> “I can’t do anything, I can’t even clean or anything. When this pain hits me, it hits me to a point that I just sleep. I must sleep because the pain runs like straight to my heart. Because when it strikes like I struggle to breath. The pain is so overwhelming I can’t do anything.” **(A014- Woman-living-with-HIV in her 20s)**

> “There is nothing I can do because it starts in the foot and then when it pains, I can’t move because that pain is very hot [painful], I must stay and feel it until it stops.” **(J030- Man-living-with-HIV in his 50s)**

### Theme 4: Management of pain

This theme describes different ways participants used to relieve their pain. These included alleviating the pain through pharmacological and non-pharmacological means.

#### Pharmacological treatment through pain relief medication

Most participants, both men and women, reported using pain relief medication such as paracetamol, Ibuprofen and analgesic gels to alleviate the pain experienced.

#### Non-pharmacological strategies

Participants in this study also indicated alternative ways of relieving the pain through non-pharmacological practices including resting the body by sleeping, walking, and soaking in warm water. Two interesting cases were noted during the analysis in which participants described unconventional ways of treating pain including physical harm and drinking lots of water.

> “When that pain starts in my chest, I tell my child to beat me here, at the back ‘cause that pain … When he beats me here at least, it’s like he massages. He beat that pain, and after that it’s going to be normal…ja. When that pain starts, I go to the wall and beat myself with my back…you see. It goes down, the pain…Yes. I can sleep but if I don’t do that, ja I won’t sleep.” **(J011- Male in his 30s)**

> “There are some days it hurts a lot, but I can walk around although it hurts. When it hurts a lot I drink a lot of water, I drink a lot of water and mix it with lemon, and it gets a bit better.” **(A074- Female in her 20s)**

Another participant shared that she consulted with a traditional healer for pain treatment.

> “I went to traditional healer, but still….and then slowly they gave me something to rub whenever I feel the pain. I would rub it but then still, nothing changed.” **(A106- Female in her 20s)**

### Theme 5: Patterns of pain disclosure

Participants’ disclosure about their pain varied from full disclosure of pain, selective or partial disclosure to non-disclosure of pain. Most participants had reported their pain to at least some people close to them. Patterns of disclosure did not differ between those living with and without HIV.

#### Full disclosure of pain

There were a few instances of full disclosure, where participants, both male and female, had told all the key people in their life including, family, friends, colleagues and health care providers.

> “Everybody knows. My [parent], I have another [sibling], they are not staying with us. I have two [siblings], we are three. So, they are not staying with us. But I told them everything about the pain. Even my neighbour. I told my neighbour.” **(A047- Female in her 30s)**

> “All of them…My [parent], my [siblings] they know …Even my [other sibling], even [though] they’re not staying with us…Uh. Yes, she [wife] knows. Even my child knows.” **(J011- Male in his 30s)**

What was more common was a certain cautiousness based on who participants decided to talk to about their pain and how much they decided to tell. Some participants chose only to share the information about the pain with some people (selective disclosure) or reserved how much they shared about the pain (partial disclosure).

#### Selective disclosure of pain

For some participants, their decision to tell some people and not others, was based on trust. This style of disclosure was only reported by people living with both chronic pain and HIV. One of the male participants shared his reason for deciding not to tell some of his friends. He said that:

> “You see, it’s like they are my friends but there are things I won’t be able to share with them. Like I share with this guy [the trusted friend], that’s how it is. You won’t trust all your friends.” (**J013-Man-living-with-HIV in his 30s**)

In addition to trust, participants living with both chronic pain and HIV selectively disclosed to manage HIV stigma. One of the female participants had told her sibling, children, neighbour, two friends, her boss and colleagues, including how much pain she is in. She doesn’t tell “each and every one” however, because they might think she is HIV positive. She said:

> “…. because people would be surprised that I have the pain. ‘Does she have HIV?’ asking things like that. Others do not accept this well.” (**A096-Woman-living-with-HIV in her 40s)**

#### Partial disclosure of pain

Other participants disclosed their pain to people close to them; however, they either didn’t share how severe the pain was, or didn’t keep them updated about painful episodes. There were several reasons for this, including: not wanting to stress their family members, friends or colleagues and to some extent retaining the agency for how to deal with the pain episode.

Participant A013 reported that she chose not to tell her children when she had severe pain episodes to keep them from stressing.

> “I don’t want to stress them [the children] a lot because I’m getting sick. You can see they are getting stressed. That does prevent … that makes me not tell them [about the pain severity or episodes of pain] because most of the time when I’m telling them, you can see that eish! … they are not relieved. Even uh, at school they called me one day … I was sick with this back pain. I couldn’t walk. They told me that uh, my first born. She was just sitting. She couldn’t do anything. So that is why I don’t want to tell them. Because they won’t concentrate at school.” **(A013- Woman-living-with-HIV in her 40s)**

Another participant chose to not tell her parent when she was experiencing pain, to keep them from forcing her to go to the clinic. She said that:

> “I didn’t tell her it hurts how much because… she would’ve said go to the clinic by force, so I told her it hurts only.” **(A074- Female in her 20s**)

#### Non-disclosure of pain

There was only one participant who had not told anyone about her pain. Her reasons included a combination of the ideas already expressed above:

> “I don’t have parents, so, I’m left with only my [siblings], but they live very far. But letting my grandparent know about this, they are gonna be very heartbroken and they are very sensitive. They… are over a hundred and imagine telling them such a thing. Their heart is very fragile… I’m afraid to tell them cause if I tell them, it’s gonna kill them.” **(A098- Female in her 20s)**

### Theme 6: Reasons for disclosure of pain

Participants shared their reasons for telling people about their pain. Their reasons ranged from: disclosing to recruit social support, transparency and in case of emergency, and to avoid judgment by others around them. To recruit social support Recruitment of social support described by participants included practical support and financial support. Women tended to recruit practical support whilst men tended to ask for financial support.

One of the participants shared that she told her neighbour about the pain so that the neighbour could help:

> “So that when I am not feeling alright, she (Neighbour) must help my child, so that she can help my child. I stay next [to] her.” **(A014- Woman-living-with-HIV in her 20s**)

Other women described telling their family and friends when a severe pain episode occurred. The family and friends would then arrive to help them bathe and dress before taking them to the clinic or hospital, and help with household chores.

> “I call my friend when I am in pain. She will take the phone and call my child or sometimes call my siblings to tell them I am not well. She will then take water to bath me, put my clothes on and do everything. After all that they (family and friend) take me to the hospital, she (friend) cleans at my house, and does my washing.” **(A084- Woman-living-with-HIV in her 40s)**

A male participant described disclosing to his family about his pain to try and get financial support.

> “Uuuh, [the] reason I tell [told] them [is because] I was needing help. Like as you know, I’ve been injured years back. I need help to go to private hospital and then help me to do an operation and check what is it inside this hand because I do feel pain always since I’ve done the operation at the public hospital. So maybe they can help me. Private hospital is the one that can help because they are much [more] concerned.” **(J007- Male in his 30s)**

The attempt by this man was unsuccessful, however. He said that:

> “But as I told them, nobody [in the family] was concerned… They all said, ‘You must be on your own brother’. [No one] seems to care about that because there was not any response since then.” **(J007- Male in his 30s)**

#### Transparency and in case of emergency

Several of the participants expressed the need for transparency reflected in their prerogative that people close to them ‘must know’ about their pain and in anticipation of a severe pain episode so they could help.

One of the participants shared that:

> “Because we live together. I don’t hide anything from her. I want her to know so that even when she wakes up and I have passed on, when they ask her, she can say I told her about the pain.” (**J064- Man-living-with-HIV in his 40s)**

Another participant expressed that people close to her must know in case of emergency so they would know what to do. She said that:

> “Unless maybe something can happen, they know that I told them about the pain…They will know what to do, maybe they can call for the ambulance or they can take me to the doctor or something or hospital.” **(A005- Female in her 40s)**

#### Avoiding being judged as lazy

A few participants from both men and women interviews reported anticipated negative judgement from others around them. Participants shared that they chose to tell people around them about their pain to avoid the possibility of being judged as lazy.

> “Ja. I told them because they started to see … to them I was reflecting I was lazy. So, they started to ask “Why … what is the problem with you? Why don’t you do this and this?” Then I told them ..I have a scoliosis problem” **(J088- Male in his 20s)**

> “I wanted them to understand what I was going through, that way they don’t get angry at me and assume that I am lazy, and I don’t want to work. They have to understand that there are some things I can’t do when I’m sick.” **(A047- Female in her 30s)**

## DISCUSSION

In this qualitative study of 34 individuals living with chronic pain in rural South Africa, we explored how chronic pain was understood and experienced, how chronic pain affected activity levels, patterns around pain disclosure and how disclosure influences recruitment of social support. So we could understand the contributions of HIV to the experience of pain, our cohort was made up half of individuals living with HIV, and half without. In participants with HIV, living with HIV was described as preferable to living with pain. As expected, participants described persisting with activity but the reasons across both those living with and without HIV were economic concerns, fear of stigma or a lack of support. Participants typically did disclose their pain to important people in their lives but they were selective about who they told, and how much. Women commonly used disclosure to recruit social support whereas this was not common in men. Patterns of activity, disclosure, and type of support recruited did not differ between those with and without HIV.

Quantitative pain severity and interference scores were similar to our other South African cohorts [10,11]. Participants described pain that was persistent but also with unpredictable episodes of severe pain. Emotions reported in response to the pain were hope that it would resolve, or fear about what it meant for the future, fearing disability and the unknown. These emotions were similarly recorded by a Ghanaian cohort with chronic low back pain [30] who also reported frustration and distress. A meta-ethnography of qualitative studies from the global north, revealed different emotional responses to the pain, however. Participants in those studies reported feeling worthless, afraid, agitated, ashamed, and guilty as they grappled between culturally-accepted biomedical understandings of pain and their own experiences [31,32]. These geographical differences reinforce the need for local studies to understand the experience of pain.

There were a variety of perceived causes of pain including following surgery, illness or accidents. Trauma and accidents as perceived causes of pain are not surprising considering the cohort was young (mean age 37). Additional perceived causes of pain included stress or ‘thinking too much’, an idiom for psychological distress or depression [33] with nearly half the cohort reporting some depressive symptoms. Participants also described non-western biomedical understandings of pain such as altered physiology (“I no longer breathe using my lungs, but I breathe using my heart so that’s why after some time it tires.”) and pain caused by witchcraft, which is a common interpretation of pain in South Africa [34,35]. Only one participant in this study spontaneously reported consulting a traditional healer to treat pain but this practice is common in South Africa, particularly in South Africans with the lowest socioeconomic status [36]. These data suggest that a trained healthcare professional or peer leader from the community who has the intercultural competence to combine both Western physiological and local understandings of pain may be most appropriate to run a pain management programme in the rural South African context [37].

The study context involved frequent manual labour as the norm, driven by the cohort’s low educational status, which led to predominantly manual occupations, and the rural setting, where tasks like walking to collect water were common. Similar to our previous quantitative studies [10,11], the qualitative data here showed participants’ persistence with activity, and this pattern was present both in those living with and without HIV. Our previous hypothesis was that PLWH and chronic pain would persist with activity to conceal their HIV status [10], but only one woman living with HIV and chronic pain reported doing so here. The most commonly reported reason for persistence in those living both with and without HIV, was economic, with fear of losing income or even their job if they were absent. These qualitative findings align with the quantitative data, which showed that economic concerns predicted objective activity levels [10]. Additionally, anecdotal evidence from another pain study of South African men indicated that fear of income or work loss interfered with attendance at a pain management programme [18]. As economic concerns driving persistence with activity despite pain was also described by participants without HIV, this suggests intersectionality of poverty and chronic pain.

In our previous actigraphy study, 10% of people had not told anyone in their life about their pain and this seemed to be due to fear of discrimination for having HIV [10]. We wondered how such a lack of disclosure would impact on recruiting social support for assistance. In this study of 34 individuals, only one person had not told anyone about their pain, and she was living without HIV. Almost all the participants, male and female, had told select people; the men described choosing people they could trust, and women framing it as avoiding people who gossiped, because stigma, either HIV or social (e.g. being perceived as lazy), was present. In those people that participants did share with, sometimes participants only partially shared the intensity or extent of their pain so that they could retain their agency or so the other would not worry too much. Indeed, what emerged was a dance of communication: to share information but avoid being the brunt of ‘gossip’ (social or HIV stigma), to recruit support but not lose agency, to share with people in their life but not stress those same people too much.

Previous qualitative data from rural South Africa suggests that it is culturally acceptable for women to disclose their pain to recruit support [34]. Women here described social support in terms of elaborate rituals of care from family, friends and neighbours who would come in and take over including housework, bathing the participant and taking them for medical care. Similar support from family and neighbours was described in women living with painful rheumatoid arthritis in Soweto, an urban township near Johannesburg [38] but not the same extensive rituals of support as reported here, such as participants being bathed. Rituals of support have been reported elsewhere in Africa where Ghanaian family members would use ointment to massage the backs of participants who were male and female physiotherapy patients with chronic low back pain [30]. Individualism versus collectivism (like the culture here in rural South Africa) moderates the relationship between fear avoidance and pain intensity [39] and so it would it would be interesting to explore in a future study whether these care-behaviours were beneficial to individuals living with pain or reduced self-efficacy and promoted fear avoidance.

Men in this study were more open about sharing their pain with others compared to other rural South African cohorts, where men expressing pain was highly taboo [34]. There may have been some bias, as more open men may have been more likely to volunteer to participate in a qualitative study. That said, men disclosed their pain with others to avoid being seen as lazy (another kind of social stigma) and to request financial support, but did not recruit practical support like the women did, or have it offered. This different pattern of support (and persistence with activity) likely speaks to the gender role identity of being male, which was clearly articulated in another study of South African men living with chronic pain [40].

There were limitations to our study. The data were collected from one site, in one province in South Africa, and so the findings may not be generalisable to other areas in South Africa. Indeed, this population was involved in a ten-year longitudinal study and we have found elsewhere that individuals with pain who are involved in research pain studies tend to improve, possibly as they believe someone cares [16,41]. Severity of depressive symptoms was much less than in our quantitative studies [10,11,42]. Indeed, half the cohort here had no depressive symptoms, and another one-fifth only minimal symptoms. As depression is associated with lower activity [43], the cohort here may have been more active than our other cohorts but we cannot say with certainty as we did not measure physical activity objectively in this cohort. As subjective functional interference and objectively measured activity did not correlate in our previous cohort of individuals living with chronic pain and HIV [10], and pain intensity has similarly failed to correlated well with objective activity in other parts of the world [44], comparing the qualitative responses with objective actigraphy data would have been informative. Although the interviewers (AR and JM) are Black South Africans, their home languages are TshiVenda and Xitsonga respectively. They can understand isiZulu and dialects of Sotho but ultimately interviews ended up being conducted in English. It would have been ideal to interview recipients in their home language as some of the nuance of participants’ experiences may have been lost. Whilst the males in the study were comfortable conversing in English, their second language, this was not always the case for the women. As such, we recruited support from an interpreter in the clinic who was not research trained. We only used this interpreter for two interviews before deciding to recruit women who were comfortable conversing in English, and whilst this improved communication, it may have introduced bias in including women who were more acculturated to the English language and culture.

In this context of high HIV and poverty, participants persisted with physical activity most of the time primarily because economic concerns gave them little choice. Pain disclosure to selected others was used to recruit social support during episodes of severe pain in women. Support requested by men was financial, if they did ask for help at all. There were very few differences between the experiences of participants living with HIV and those without, suggesting intersectionality between pain and poverty rather than pain and HIV. Within this high HIV prevalence context, having HIV did not seem important in relation to how pain is managed. Rather, our findings suggest that activity levels, disclosure and recruitment of support in people living with chronic pain are influenced by low income and gender.

## Data Availability

All data produced in the present study are available upon reasonable request to the authors.

## Acknowledgements

We are grateful to the Ndlovu Care Group for allowing us to collect data at their Research Centre including the help and support of the clinic staff. In particular, we are grateful to Stephina Molepa and Victoria Molepa. We are grateful too to the participants for the sharing of their stories and experiences.

## Author contributions

MM and AW conceived the study design, oversaw data acquisition, contributed to data analysis and interpretation, and wrote the draft manuscript. RP, TP, ME conceived the study design and contributed to data interpretation. AR and JM acquired, analysed and interpreted the data. TR contributed significantly to data analysis and interpretation. CNU and HT commented on study design and contributed to data acquisition. All authors critically reviewed the work and approved the final version.

## Data statement

Qualitative data are available upon reasonable request from the senior author.

## STRENGTHS AND LIMITATIONS OF THIS STUDY

The qualitative design helped explore a phenomena of maintained levels of physical activity whilst living with chronic pain, reported in quantitative studies of poor South Africans.

- The recruitment of a sample half living with HIV and half without, helped explore the contribution of HIV to the experience and impact of chronic pain.
- This is the first qualitative study exploring pain disclosure and social support in people living with chronic pain in South Africa.
- Data were collected from one site in one province in South Africa and so may not be generalisable to all rural areas in South Africa.

## Footnotes

1 This refers to the participant boiling the water and then placing into a bath for the children.

